# Multi-class classification of central and non-central geographic atrophy using Optical Coherence Tomography

**DOI:** 10.1101/2025.05.27.25328446

**Authors:** Sadia Siraz, Hindolo Kamanda, Sina Gholami, Ahammed Sakir Nabil, Sally Shin Yee Ong, Minhaj Nur Alam

## Abstract

**Purpose:** To develop and validate deep learning (DL)-based models for classifying geographic atrophy (GA) subtypes using Optical Coherence Tomography (OCT) scans across four clinical classification tasks.

**Design:** Retrospective comparative study evaluating three DL architectures on OCT data with two experimental approaches.

**Subjects:** 455 OCT volumes (258 Central GA [CGA], 74 Non-Central GA [NCGA], 123 no GA [NGA]) from 104 patients at Atrium Health Wake Forest Baptist. For GA versus age-related macular degeneration (AMD) classification, we supplemented our dataset with AMD cases from four public repositories.

**Methods:** We implemented ResNet50, MobileNetV2, and Vision Transformer (ViT-B/16) architectures using two approaches: (1) utilizing all B-scans within each OCT volume and (2) selectively using B-scans containing foveal regions. Models were trained using transfer learning, standardized data augmentation, and patient-level data splitting (70:15:15 ratio) for training, validation, and testing.

**Main Outcome Measures:** Area under the receiver operating characteristic curve (AUC-ROC), F1 score, and accuracy for each classification task (CGA vs. NCGA, CGA vs. NCGA vs. NGA, GA vs. NGA, and GA vs. other forms of AMD).

**Results:** ViT-B/16 consistently outperformed other architectures across all classification tasks. For CGA versus NCGA classification, ViT-B/16 achieved an AUC-ROC of 0.728±0.083 and accuracy of 0.831±0.006 using selective B-scans. In GA versus NGA classification, ViT-B/16 attained an AUC-ROC of 0.950±0.002 and accuracy of 0.873±0.012 with selective B-scans. All models demonstrated exceptional performance in distinguishing GA from other AMD forms (AUC-ROC>0.998). For multi-class classification, ViT-B/16 achieved an AUC-ROC of 0.873±0.003 and accuracy of 0.751±0.002 using selective B-scans.

**Conclusions:** Our DL approach successfully classifies GA subtypes with clinically relevant accuracy. ViT-B/16 demonstrates superior performance due to its ability to capture spatial relationships between atrophic regions and the foveal center. Focusing on B-scans containing foveal regions improved diagnostic accuracy while reducing computational requirements, better aligning with clinical practice workflows.

## Introduction

Age-related macular degeneration (AMD) is one of the most common causes of vision impairment in developed nations, particularly affecting individuals over 65 years old^1^, with global prevalence expected to reach 288 million by 2040.^2,3^ AMD has three different stages - early, intermediate, and advanced. Geographic atrophy (GA) is characterized by the later stage of dry AMD causing progressive, irreversible degeneration of the retinal pigment epithelium (RPE), overlying photoreceptors (PRs), and underlying choriocapillaris in the macula.^4^ GA usually begins in the parafoveal region, called (i.e. non-central GA, NCGA), and can progressively extend to the fovea (Central GA, CGA) over several years.^5^ However, in some rare cases, central fovea can be affected at the disease onset. Atrophic regions are strongly associated with dense scotoma, leading to significant visual impairment. Consequently, CGA is often accompanied by severe reductions in central visual acuity. Even when atrophy remains non-central, declines in functional vision can substantially impact essential tasks such as reading and facial recognition.^6^ Therefore, the task of distinguishing cases of AMD with and without GA, and central or non-central GA is essential for earlier intervention strategies, personalized treatment approaches, and improved monitoring of disease progression in clinical trials. However, the classification between CGA and NCGA can be challenging primarily due to the subjective nature of defining the exact boundary of the central area on the retina. This can vary depending on the imaging modality used, the observer’s interpretation, and the presence of subtle changes in the macular anatomy, leading to inconsistent diagnosis across different clinicians and studies.

Optical coherence tomography (OCT) and fundus autofluorescence (FAF) are the most widely utilized imaging modalities to monitor GA development and progression.^7^ However, OCT provides a superior, high-resolution, cross-sectional visualization of GA lesions. It offers critical depth resolved structural insights that surpass the capabilities of FAF and it is recognized by the Classification of Atrophy Meetings (CAM) group.^8^ Artificial Intelligence (AI) based models have been successful when dealing with a large number of individual B-scans in an OCT volume.^9^ These models excel at identifying subtle patterns in retinal structure that may exclude the need for human graders. Some of the recent studies have shown remarkable accuracy in automated algorithms developed for GA lesion segmentation.^7,9,10^

Despite these advances, a significant research gap exists in GA classification because of its phenotypic presentation and anatomical location. Different subtypes of GA represent a critical distinction with direct relevance to clinical practice and research. Central involvement leads to significant loss of visual acuity, whereas in NCGA, central vision could still be preserved for a longer time, even though there may be paracentral scotomas. Additionally, distinguishing GA from non-atrophic AMD and a healthy retina is crucial for early detection and monitoring of patients who may be at risk of developing atrophy. To date, no comprehensive deep learning (DL)-based approach has been developed specifically for the multi-class classification of these GA subtypes (especially CGA vs NCGA).

In this study, we address this critical knowledge gap by developing and validating DL-based models using OCT scans for four clinically relevant classification tasks using a clinical dataset: i) distinguishing CGA from NCGA; ii) multi-class classification of CGA vs. NCGA vs. no GA (NGA); iii) binary classification of GA (any type) vs. NGA; and iv) differential diagnosis between GA and other forms of AMD. For each of the cases, we adapted two distinct experimental approaches - i) utilizing all B-scans within each OCT volume to capture comprehensive pathological features, and ii) leveraging the B-scans of interest that include foveal regions and crucial areas exhibiting distinctive GA characteristics within each OCT volume. Using only the B-scans of interest is helpful in improving diagnostic accuracy while reducing computational burden compared to processing entire OCT volumes. Our OCT based GA models can provide ophthalmologists with automated tools for rapid GA phenotyping, enabling personalized management strategies based on specific GA subtypes and improving patient selection for emerging therapies. This is particularly of clinical and scientific interest due to the recent clinical trials of FDA approved drugs for GA prevention.

## Methodology

This section provides the description of the step by step methodology that has been adopted for the completion of GA classification tasks. We have mainly conducted two sets of experiments as already mentioned before *i) Case 1*-leveraging complete OCT volumes, incorporating all B-scans to capture the full spectrum of retinal pathology for different visits of a patient, *ii) Case 2*-selecting B-scans which include the foveal region from an OCT volume and discarding others. The foveal area represents the most functionally significant portion of the macula, responsible for central vision and fine detail discrimination. When evaluating GA progression, foveal B-scans provide critical information about RPE integrity, outer retinal layer preservation, and choroidal transmission changes that characterize atrophic regions. Moreover, the phenomenon of foveal sparing, where atrophy surrounds but does not yet involve the central fovea, has significant prognostic implications for visual function. It can only be properly assessed through targeted foveal imaging. Ophthalmologists rely on these specific B-scans to make treatment decisions and monitor disease progression, as even small changes in foveal involvement can dramatically impact a patient’s visual experience and quality of life. Thus adopting the approach of case 2 has significantly increased our computational efficiency. The rest of the steps adopted for the classification tasks for both of the experiments are the same as shown in figure 1.

**Figure 1:**
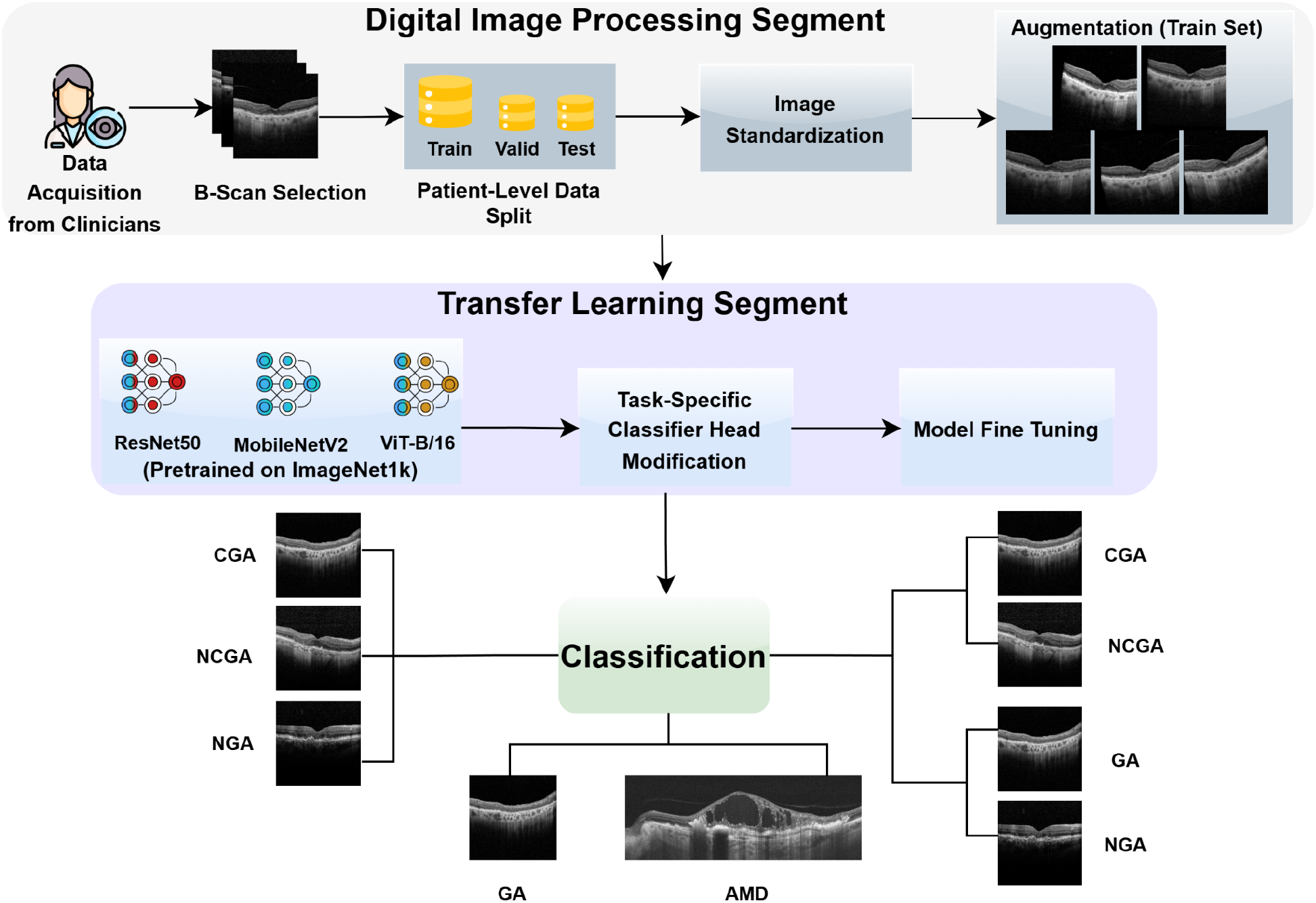
Step by step methodology of DL based GA classification task-Digital Image processing segment; Transfer learning segment; Classification task. DL = Deep Learning; GA = Geographic Atrophy; CGA = Central GA; NCGA = Non-Central GA; NGA= No GA; AMD = Age-related Macular Degeneration.

## Data

### Data Acquisition

We utilized OCT scans collected from two distinct sources to train and evaluate our DL models. For our primary classification tasks (CGA vs. NCGA, CGA vs. NCGA vs. NGA, and GA vs. NGA), we employed a private clinical dataset from Atrium Health Wake Forest Baptist. For the differential diagnosis task between GA and other forms of AMD, we supplemented our private GA data with 4 publicly available datasets DS1-DS4.^11–14^ This study was approved by the Institutional Review Board of Atrium Health Wake Forest Baptist and adhered to the principles of the Declaration of Helsinki.

#### Wake Forest Dataset

A retrospective chart review, including records from Jan 1, 2013 to Jan 28, 2023, was conducted. Charts were identified using the ICD9 codes 362.50 and 362.51 and ICD10 codes H35.30 and H35.31. Patients were excluded if they : 1) lacked sequential OCT imaging prior to subfoveal conversion; (2) presented with subfoveal involvement at initial consultation; (3) exhibited significant confounding macular pathologies, including choroidal neovascularization (CNV), diabetic macular edema (DME), or retinal vein occlusions (CRVO/BRVO); (4) demonstrated neovascular AMD; or (5) had insufficient longitudinal data. Additionally, patients who had received treatment for retinal conditions that might affect imaging interpretation were excluded. Following this comprehensive selection process, our final cohort consisted of 104 patients who had adequate imaging data for our analysis. All OCT scans were graded by an expert grader at Atrium Health Wake Forest Baptist. CGA was defined as GA involving the central 1mm ETDRS circle, while NCGA was defined as GA outside the central 1mm ETDRS circle. From these 104 patients, we incorporated appropriate OCT volumes for each classification category, 258 OCT volumes for CGA, 74 volumes for NCGA and 123 OCT volumes were graded as NGA. For each patient, we included OCT scans obtained during multiple clinical visits, capturing disease states at different time points. Table 1 represents the demographic features of the selected patients.

**Table 1:**
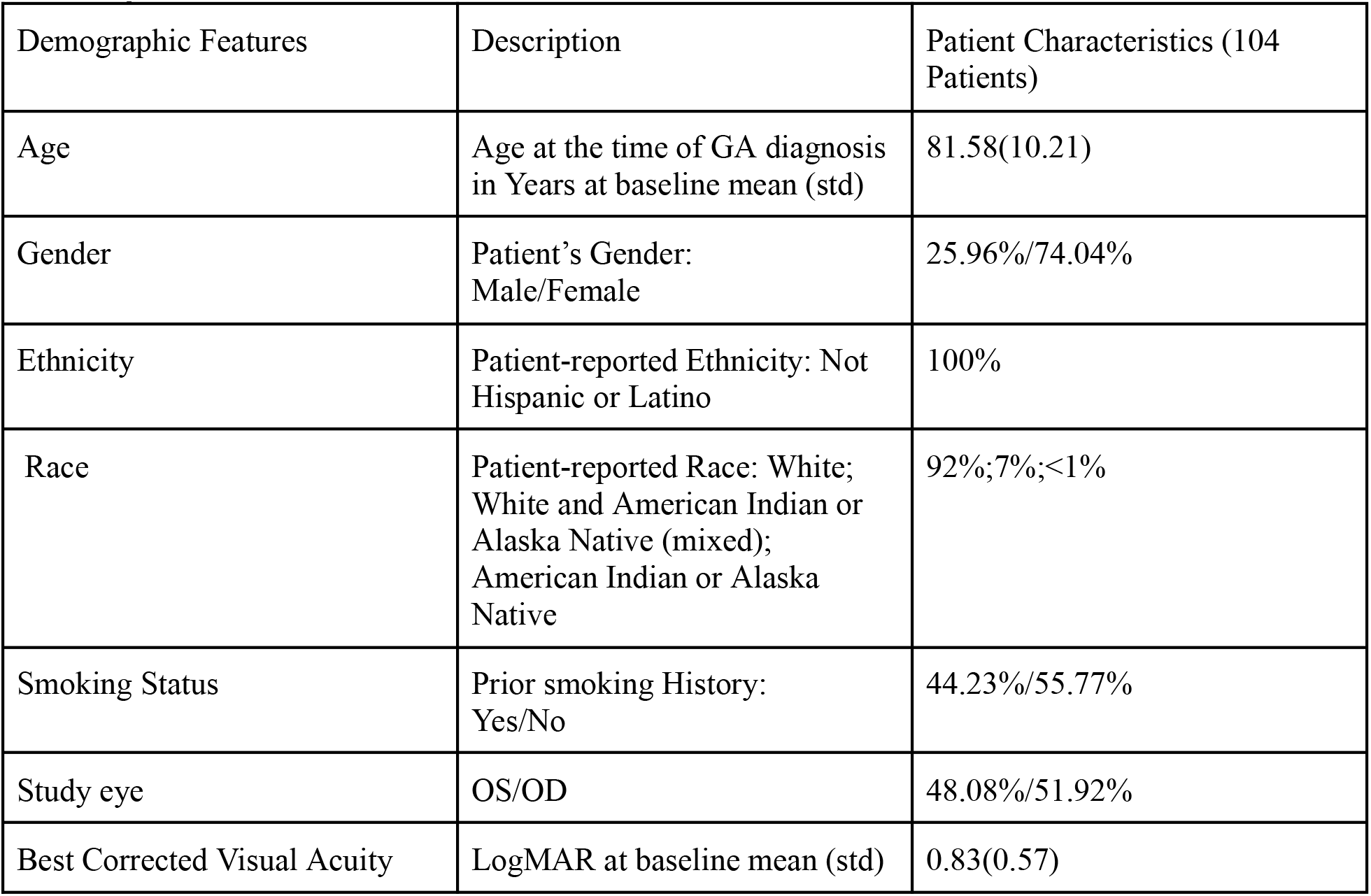
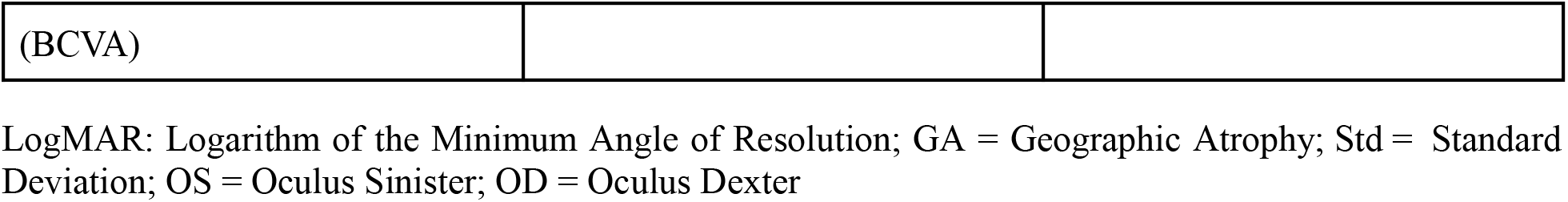
List of demographic and visual factors considered in our analysis for Atrium Health Wake forest Baptist Dataset.

The OCT volumes were acquired using Heidelberg HRA+OCT Spectralis Version 2.5.9 (Build 2048) devices with Heidelberg Heyex software. Most patients were not fully dilated at the time of imaging, reflecting real-world clinical conditions. Each volume had a resolution of 512 × 496 × 25 voxels with 25 B-scans per volume, covering approximately 5.9 × 1.9 × 0.25 mm^3^ of retinal area. This scan density provided comprehensive visualization of the macular region, allowing for detailed assessment of GA features.

#### Public dataset (DS1-DS4)

For our GA vs AMD classification task, we leveraged AMD data from four publicly available OCT datasets:

1. DS1: We selected AMD cases from this dataset, which originally contained retinal images from 45 subjects evenly distributed across Normal, AMD, and DME categories (15 subjects per category)^11^.
2. DS2: From this dataset of 500 OCT images, we extracted AMD cases. The original dataset included images captured using two distinct fields of view (3-mm and 6-mm), with 3-mm files containing 304 scans per patient and 6-mm files containing 400 scans. These images were acquired using a spectral-domain OCT system with a center wavelength of 840 nm (RTVue-XR, Optovue, CA)^12^.
3. DS3: We utilized AMD cases from this open-access database containing over 500 high-resolution retinal images acquired with a Cirrus HD-OCT machine (Carl Zeiss Meditec, Inc., Dublin, CA)^13^.
4. DS4: We extracted AMD cases from the OCTDL dataset, which originally included over 2,000 OCT images from patients aged 20 to 93 years (male-to-female ratio 3:2, average age 63 years). The B-scan OCT images in this dataset were obtained using a raster scanning protocol with dynamic scan length and resolution, using the Optovue Avanti RTVue XR, with each scan centered over the fovea and evaluated by an experienced retinal specialist^14^.

By selectively utilizing AMD cases from these diverse public repositories, we assembled a comprehensive dataset for our GA vs AMD classification task, benefiting from varied imaging protocols and equipment specifications.

### Dataset Partitioning

To ensure robust model evaluation while preventing data leakage, we implemented a patient-level data splitting strategy. This approach maintained the integrity of our evaluation by ensuring that all OCT B-scans from the same patient were exclusively allocated to either the training, validation, or test sets. We extracted unique patient identifiers and used these as the basis for randomized stratification. All the approaches that have been implemented for the classification tasks follow the same data splitting methods. For both of the cases, the B-scans have been separated, maintaining a 70:15:15 ratio for training, validation, and testing respectively as illustrated in Figure 2. Figure 2 shows the distribution of images across the training, validation, and test sets for all four classification tasks in both experimental approaches.. This splitting methodology preserved the relationship between multiple B-scans from the same OCT volume, which is critical for maintaining clinical relevance in the evaluation process. The training set was used for model optimization, the validation set for hyperparameter tuning and early stopping decisions, while the test set remained completely isolated until final model evaluation to provide an unbiased assessment of model performance. Prior to feeding the images into the neural networks, all OCT B-scans were standardized through preprocessing steps for both of the cases.

**Figure 2.**
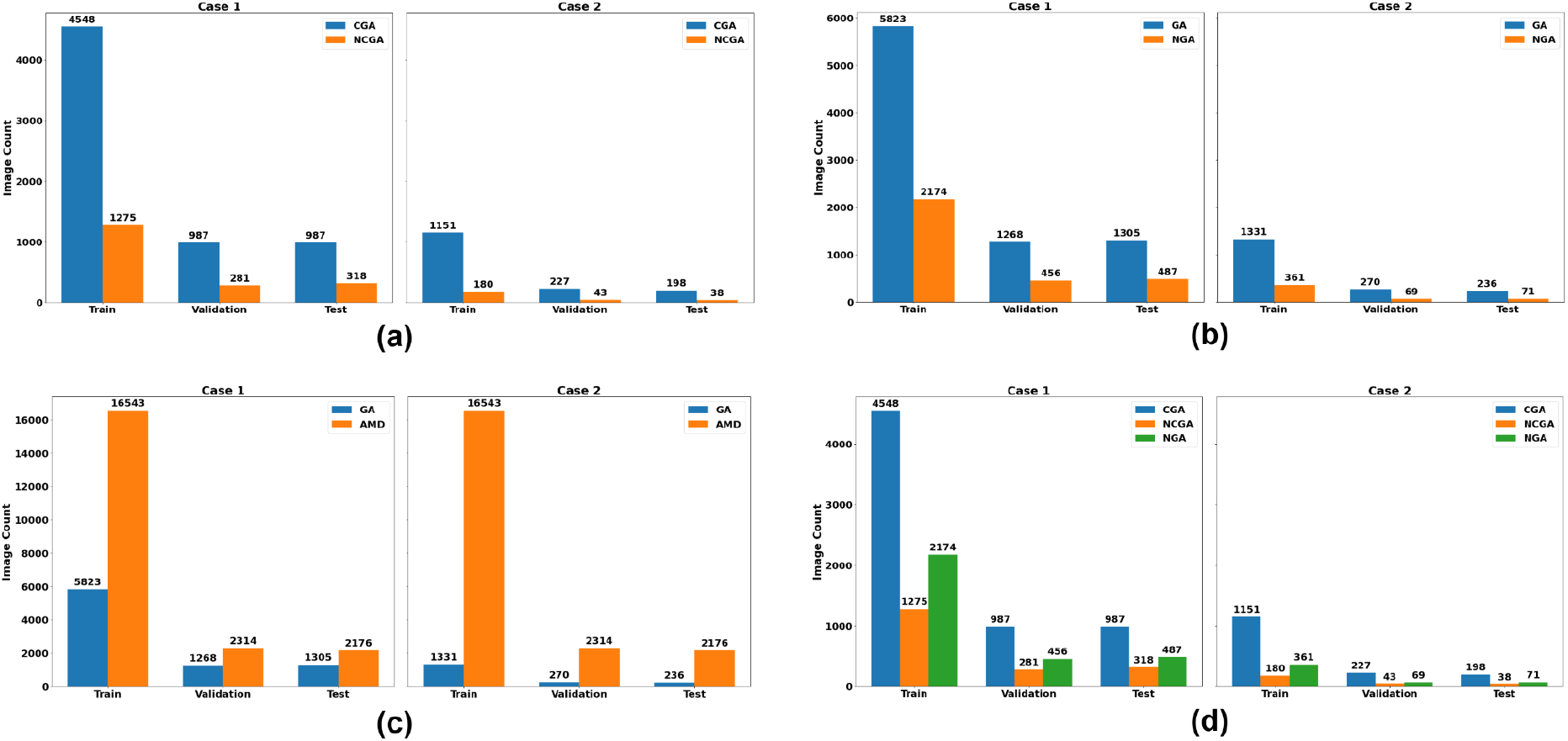
Dataset distribution across training, validation, and test sets for the four classification tasks. (a) Binary classification of CGA vs NCGA using all B-scans (Case 1) and selected B-scans of interest (Case 2). (b) Binary classification of GA vs no NGA. (c) Binary classification of GA vs other forms of AMD. (d) Multi-class classification of CGA vs NCGA vs NGA. GA = Geographic Atrophy; CGA = Central GA; NCGA = Non-Central GA; NGA= No GA; AMD = Age-related Macular Degeneration.

### Data Pre-processing

For both approaches of our experiments, the OCT B-scans underwent a standardized preprocessing sequence beginning with RGB conversion to maintain compatibility with established neural network architectures. Images were subsequently resampled to uniform dimensions (224×224 pixels) to conform to the input requirements of our selected DL models.

To enhance model robustness and generalization capabilities, we incorporated comprehensive data augmentation techniques during the training phase. These included random horizontal flipping with 50% probability, random rotations (up to 15 degrees), brightness and contrast adjustments, random affine transformations for spatial shifts, and random resized cropping maintaining at least 80% of the original content. These augmentations simulate the natural variability encountered in clinical settings and help prevent overfitting to training examples.

All images were normalized using ImageNet statistics (mean=[0.485, 0.456, 0.406], standard deviation=[0.229, 0.224, 0.225]) to leverage transfer learning benefits and accelerate model convergence on retinal pathology detection tasks.

The data handling framework incorporated dynamic batch processing with four-worker parallel loading to optimize throughput. Training data underwent the augmentation pipeline while validation and testing sets maintained deterministic processing with only resizing and normalization to ensure reproducible evaluation metrics. For all classification tasks, including binary and multi-class classification, we maintained consistent preprocessing parameters for unbiased evaluation of model architectures.

## Model Architecture

### Baseline Model Selection

For all of our classification tasks of GA for both of the cases, we have implemented and compared three well-established baseline neural network architectures: ResNet50, MobileNetV2, and Vision Transformer (ViT-B/16). Each architecture was selected for their specific advantage over others.

- ResNet50: ResNet50 is a 50 layer residual network with skip connections. It mitigates gradient vanishing issues during training, enabling deeper network training while maintaining computational efficiency. ResNet50 has demonstrated exceptional performance in medical image classification tasks due to its depth and feature extraction capabilities.^15^
- MobileNetV2: MobileNetV2 is a lightweight architecture designed for resource constrained environments. It uses depth-wise separable convolutions to reduce computational complexity while maintaining competitive accuracy. This model was included to evaluate performance-efficiency tradeoffs in clinical deployment scenarios.^16^
- Vision Transformer (ViT-B/16): ViT-B/16 is a transformer-based architecture that processes images as sequences of patches, leveraging self-attention mechanisms to capture global relationships between image regions.^17^ ViT models have shown promising results in capturing long-range dependencies in medical images, potentially beneficial for detecting subtle GA patterns across OCT B-scans.^3^

### Model Implementation

For all three architectures, we implemented a consistent transfer learning approach while making architecture-specific adaptations for optimal performance. Each model was initialized with pre-trained ImageNet weights, with weights of ResNet50’s convolutional layers frozen for robust feature extraction while only the final classification layer was modified. For MobileNetV2, we selectively froze feature extraction modules while allowing the inverted residual blocks to adapt to OCT-specific features, balancing efficiency with domain-specific adaptation. With Vision Transformer, we maintained the self-attention mechanism and patch embedding structure while adapting only the classification components to preserve its ability to capture global spatial relationships crucial for GA pattern detection. Across all architectures, we replaced the original classification layers with a custom head consisting of a 512-neuron fully connected layer with ReLU activation. Dropout of 30% was employed to prevent overfitting, and a final linear layer mapped to the appropriate number of output classes based on the specific classification task. This standardized approach to fine-tuning enabled fair comparison between architectures while the selective parameter freezing strategy optimized computational efficiency by training only task-specific components on our OCT dataset.

### Training Configuration

For both experimental cases, we standardized training parameters across architectures to ensure methodologically rigorous comparison. We employed the Adam optimizer with an initial learning rate of 1×10^−3^, selected for its adaptive learning rate properties that facilitate efficient training with sparse gradients common in DL models. To mitigate learning rate sensitivity and improve convergence, we implemented a cosine annealing scheduler that systematically reduced the learning rate to 1×10^−6^ over the training period, following established best practices for training deep neural networks. Cross-entropy loss was chosen as the objective function for all classification tasks due to its effectiveness in multi-class problems and its mathematical properties that stabilize the learning process. All models were trained for a maximum of 50 epochs, a limit established after preliminary experiments showed convergence typically occurred within this range. This training duration strikes an optimal balance between ensuring model convergence and computational efficiency, particularly important when processing the high-dimensional OCT volume data. The relatively modest epoch count was appropriate given the use of transfer learning with pre-trained model weights, which substantially reduces the required training time compared to training from scratch. Our implementation of early stopping (with a patience threshold of 20 epochs) further optimized the training process, as models would automatically terminate training when no improvement in validation accuracy was observed, preventing unnecessary computation while ensuring optimal model performance. Model selection was based on peak validation accuracy rather than training metrics to ensure optimal generalization to unseen clinical data.

All models were trained using a high-performance computing environment equipped with NVIDIA GeForce GTX 1080 Ti GPUs (11GB VRAM per GPU) running CUDA version 12.2 and NVIDIA driver 535.230.02. We employed a consistent batch size of 16 for all experiments, with models trained on individual GPUs to ensure standardized conditions across all architectures and tasks.

## Results

### Binary Classification of CGA vs. NCGA

We evaluated three DL architectures for distinguishing CGA and NCGA using OCT scans. Accurate discrimination between CGA and NCGA has significant clinical implications, as CGA typically progresses more rapidly to legal blindness and may require more intensive monitoring and early intervention strategies compared to NCGA. As shown in Table 2, Vision Transformer (ViT-B/16) demonstrated superior performance compared to both ResNet50 and MobileNetV2 across all evaluation metrics. Each model was trained five times with different random seeds to account for variability in training outcomes, and we report the mean and standard deviation of each metric. Area Under the Receiver Operating Characteristic curve (AUC-ROC) was used as the primary performance metric to evaluate our models. When utilizing all B-scans within each OCT volume (Case 1), ViT-B/16 achieved an AUC-ROC of 0.718 ± 0.002, F1 score of 0.602 ± 0.004, and accuracy of 0.780 ± 0.005. In contrast, when using only the B-scans of interest that included foveal regions (Case 2), the performance of ViT-B/16 improved for AUC-ROC to 0.728 ± 0.083. This came at the cost of a slight decrease in F1 score (0.492 ± 0.026), while maintaining comparable accuracy (0.831 ± 0.006). To evaluate the statistical significance of performance differences between architectures, we conducted paired t-tests across 5 random seeds. For the CGA vs. NCGA classification task, ViT-B/16 achieved significantly higher mean AUC-ROC compared to ResNet50 in both Case 1 (mean difference = 0.173, t(4) = −20.04, p < 0.001) and Case 2 (mean difference = 0.148, t(4) = −5.57, p < 0.01).

**Table 2:**
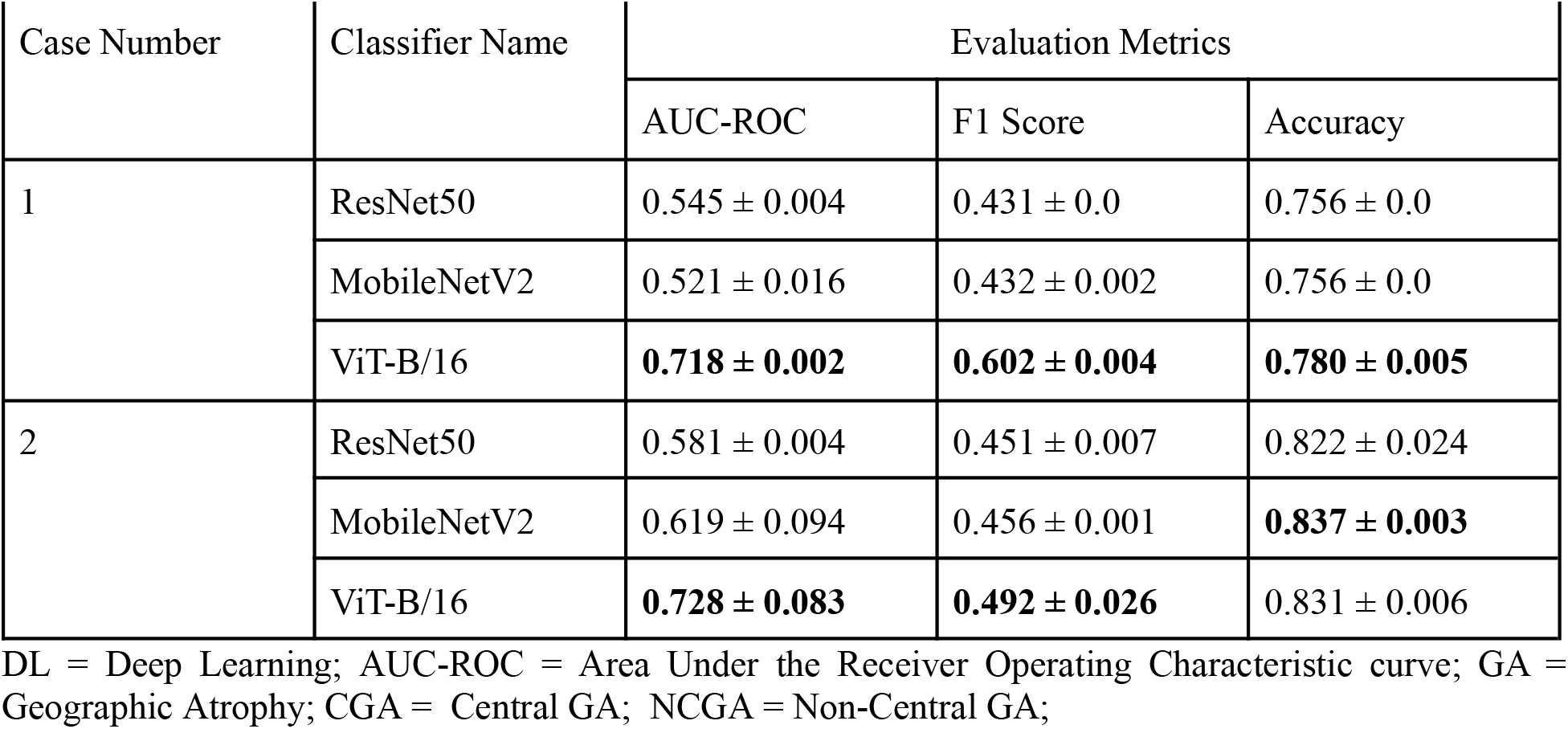
Comparative Analysis of DL Architectures for Binary Classification of CGA vs. NCGA in OCT Scans.

| Case Number | Classifier Name | Evaluation Metrics |  |  |
| --- | --- | --- | --- | --- |
|  |  | AUC-ROC | F1 Score | Accuracy |
| 1 | ResNet50 | $0.545 \pm 0.004$ | $0.431 \pm 0.0$ | $0.756 \pm 0.0$ |
| | MobileNetV2 | $0.521 \pm 0.016$ | $0.432 \pm 0.002$ | $0.756 \pm 0.0$ |
|  | ViT-B/16 | **$0.718 \pm 0.002$** | **$0.602 \pm 0.004$** | **$0.780 \pm 0.005$** |
| 2 | ResNet50 | $0.581 \pm 0.004$ | $0.451 \pm 0.007$ | $0.822 \pm 0.024$ |
| | MobileNetV2 | $0.619 \pm 0.094$ | $0.456 \pm 0.001$ | **$0.837 \pm 0.003$** |
| | ViT-B/16 | **$0.728 \pm 0.083$** | **$0.492 \pm 0.026$** | $0.831 \pm 0.006$ |

DL = Deep Learning; AUC-ROC = Area Under the Receiver Operating Characteristic curve; GA = Geographic Atrophy; CGA = Central GA; NCGA = Non-Central GA;

Notably, MobileNetV2 demonstrated the highest accuracy (0.837 ± 0.003) among all models when using only B-scans of interest, suggesting that selective B-scan analysis may enhance the performance of lighter architectures for this specific classification task.

The ROC curves in Figure 3(a-b) provide visual confirmation of the performance metrics. In Case 1 (Figure 3a), ViT-B/16 achieved the highest AUC of 0.725, displaying a curve that consistently dominates those of ResNet50 (AUC = 0.562) and MobileNetV2 (AUC = 0.539). The clear separation between the ViT-B/16 curve and the others across all threshold values indicates its superior ability to distinguish between CGA and NCGA. For Case 2 (Figure 3b), all models showed more step-like ROC curves due to the reduced dataset size, with ViT-B/16 maintaining its dominance (AUC = 0.701) over MobileNetV2 (AUC = 0.637) and ResNet50 (AUC = 0.583).

**Figure 3:**
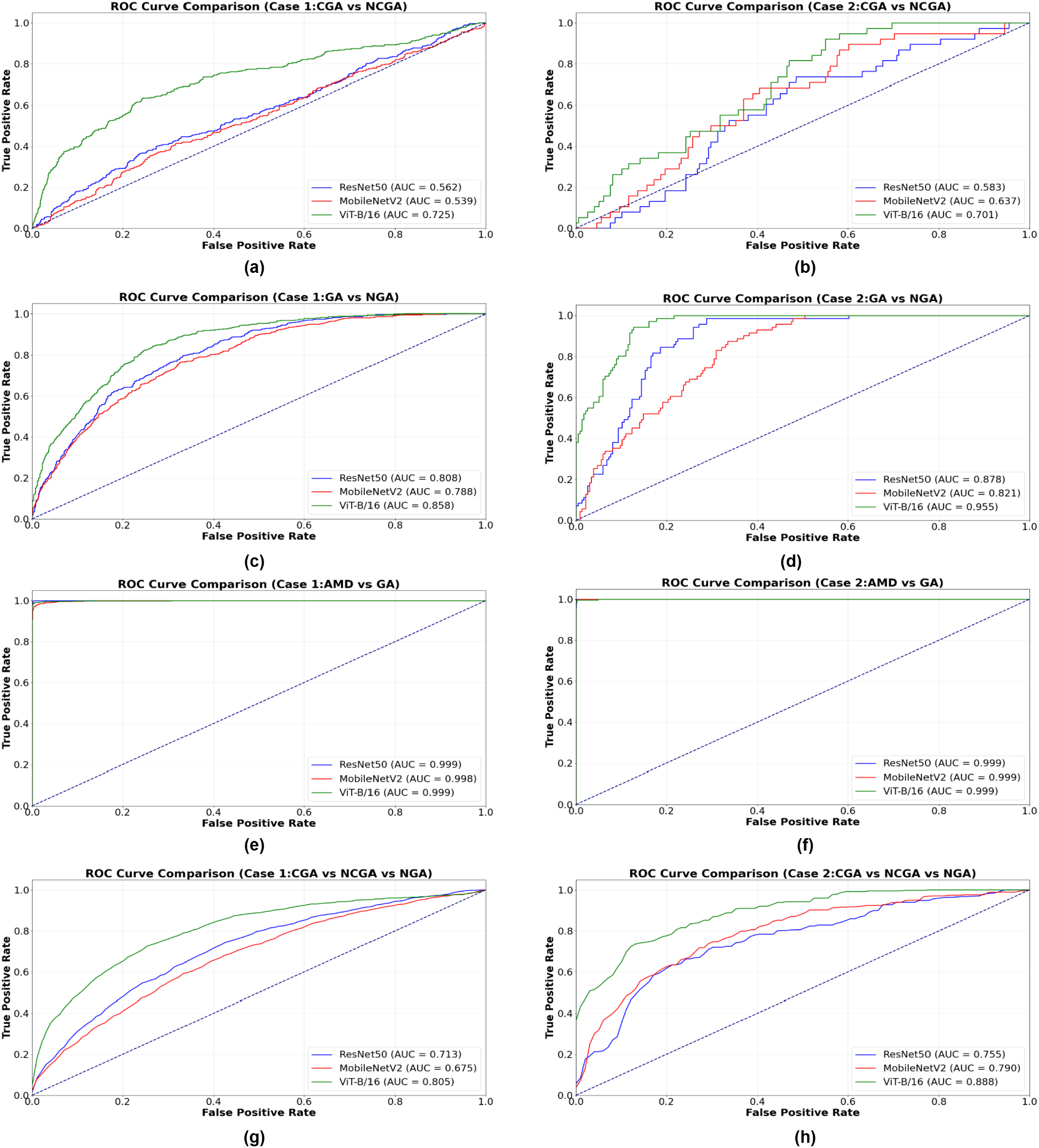
ROC Curves for different DL Models for Geographic Atrophy classification tasks. DL = Deep Learning; ROC = Receiver Operating Characteristic; GA = Geographic Atrophy; CGA = Central GA; NCGA = Non-Central GA; NGA= No GA; AMD = Age-related Macular Degeneration.

The confusion matrices in Figure 4(a-b) reveal important classification patterns. In Case 1 (Figure 4a), ViT-B/16 correctly classified 952 out of 987 CGA samples (96.5% sensitivity), but only correctly identified 77 out of 318 NCGA samples (24.2% specificity). This substantial imbalance suggests the model prioritizes CGA identification, potentially due to class imbalance in the training data. In Case 2 (Figure 4b), the model demonstrated even stronger CGA identification with 197 correct classifications out of 198 (99.5% sensitivity), but continued to struggle with NCGA detection, correctly classifying only 2 out of 38 samples.

**Figure 4:**
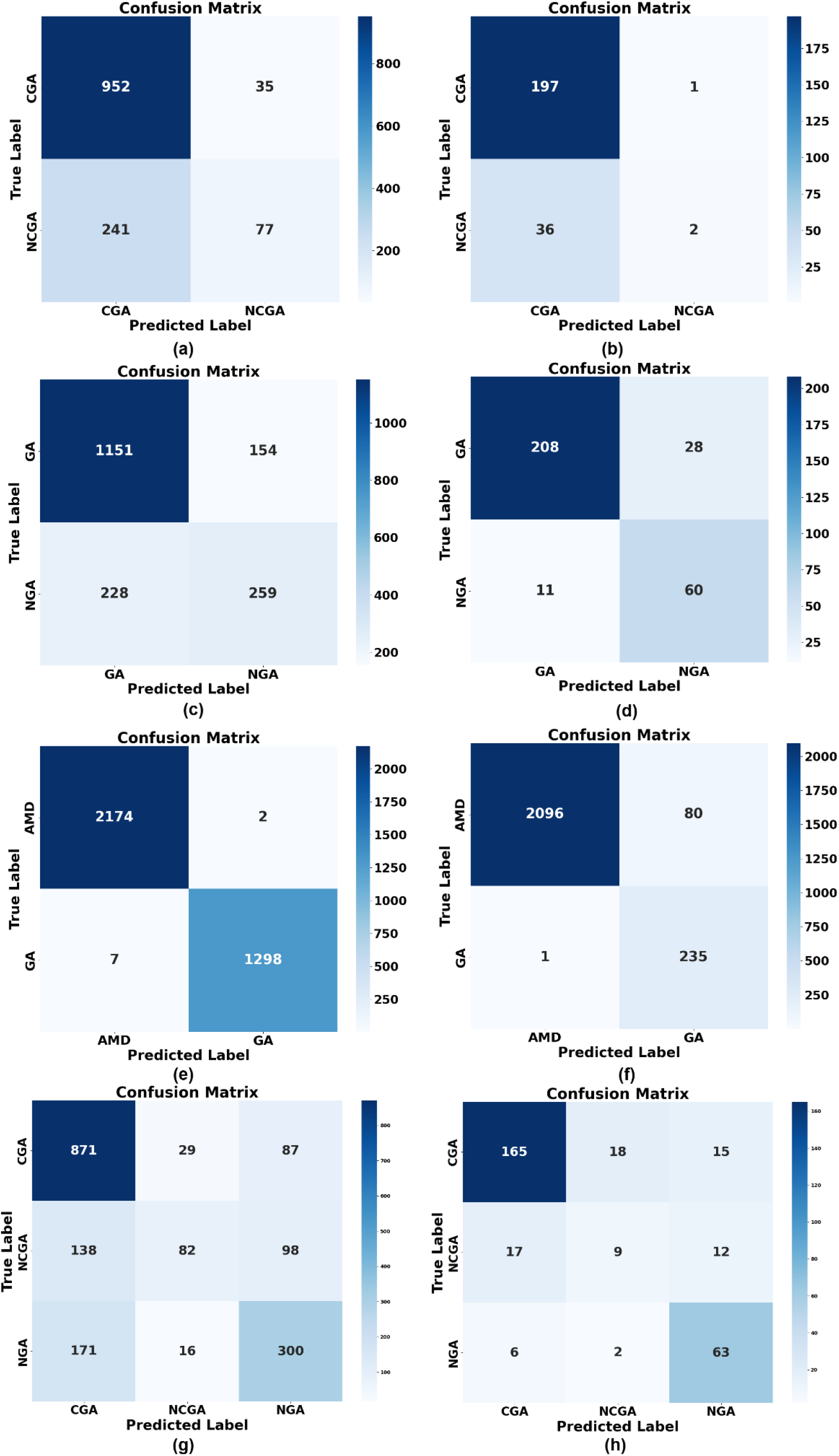
Confusion Matrix for ViT-B/16 Model-case 1- (a) CGA vs NCGA, (c)GA vs NGA (e) GA vs AMD (g) CGA vs NCGA vs NGA; Case 2- (b) CGA vs NCGA, (d)GA vs NGA (f) GA vs AMD - ResNet50 (h) CGA vs NCGA vs NGA. GA = Geographic Atrophy; CGA = Central GA; NCGA = Non-Central GA; NGA= No GA; AMD = Age-related Macular Degeneration.

### Binary Classification of GA vs. NGA

For the task of distinguishing any type of GA from NGA, ViT-B/16 consistently outperformed the other architecture across both experimental approaches. As illustrated in Table 3, when processing all B-scans (Case 1), ViT-B/16 achieved an AUC-ROC of 0.856 ± 0.003, F1 score of 0.728 ± 0.006, and accuracy of 0.791 ± 0.003.

**Table 3:**
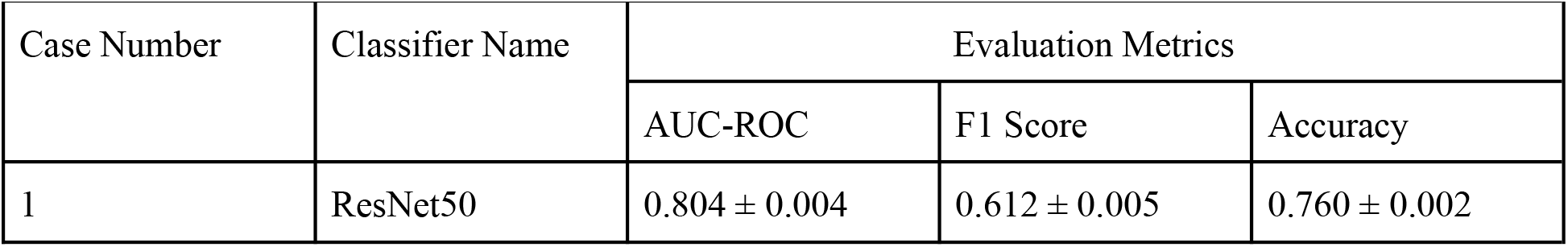
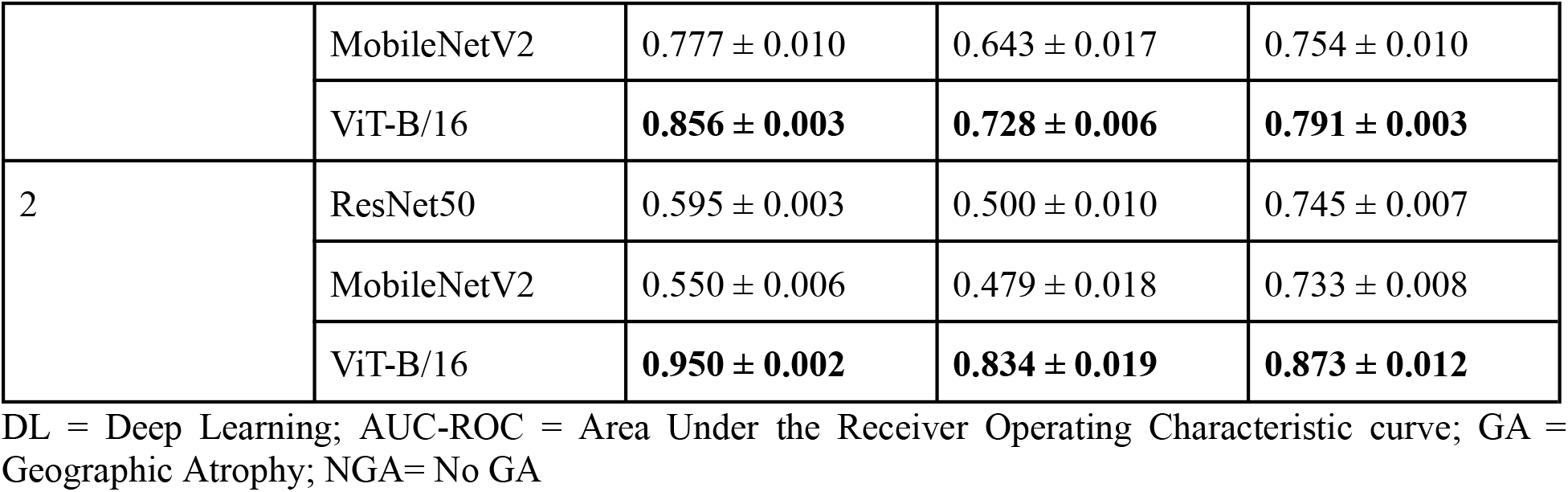
Comparative Analysis of DL Architectures for Binary Classification of GA vs. NGA in OCT Scans.

The performance improved substantially when focusing only on B-scans of interest (Case 2), with ViT-B/16 reaching an AUC-ROC of 0.950 ± 0.002, F1 score of 0.834 ± 0.019, and accuracy of 0.873 ± 0.012. This represents an 11.0% increase in AUC-ROC, 14.6% increase in F1 score, and 10.4% increase in accuracy compared to using all B-scans, highlighting the significant benefit of selective B-scan analysis for this classification task. In this classification task, ViT-B/16 significantly outperformed ResNet50 in both experimental approaches, with a modest improvement in Case 1 (mean difference = 0.048, t (4) = −13.29, p < 0.001) and a substantial gain in Case 2 (mean difference = 0.355, t (4) = −192.33, p < 0.001). The ROC curves in Figure 3(c-d) illustrate the clear advantage of ViT-B/16 for GA vs. NGA classification. In Case 1 (Figure 3c), ViT-B/16 achieved an AUC of 0.858, with a curve that shows consistently higher true positive rates across all false positive rates compared to ResNet50 (AUC = 0.808) and MobileNetV2 (AUC = 0.788). The improvement is even more pronounced in Case 2 (Figure 3d), where ViT-B/16 reaches a higher AUC of 0.955, demonstrating near-perfect performance at lower false positive rates. The steep initial rise of the ViT-B/16 curve in Case 2 indicates that the model achieves high sensitivity even at very high specificity thresholds, which is particularly valuable for clinical applications where minimizing false positives is crucial.

The confusion matrices in Figure 4(c-d) quantify these improvements. For Case 1 (Figure 4c), ViT-B/16 correctly identified 1151 out of 1305 GA samples (88.2% sensitivity) and 259 out of 487 NGA samples (53.2% specificity). In Case 2 (Figure 4d), both sensitivity and specificity improved significantly, with 208 out of 236 GA samples correctly identified (88.1% sensitivity) and 60 out of 71 NGA samples correctly classified (84.5% specificity). This 31.3% increase in specificity when using selective B-scans demonstrates that focusing on relevant regions substantially reduces false positive classifications of NGA as GA, an important consideration for clinical implementation.

### Binary Classification of GA vs. AMD

All three DL architectures demonstrated exceptional performance in distinguishing GA from other forms of AMD, as presented in Table 4. Using all B-scans (Case 1), ResNet50 and MobileNetV2 both achieved perfect AUC-ROC scores of 0.999 ± 0.0, with ViT-B/16 performing similarly at 0.998 ± 0.003. ResNet50 showed the highest F1 score (0.992 ± 0.002) and accuracy (0.995 ± 0.001) for this case.

**Table 4:** Comparative Analysis of DL Architectures for Binary Classification of GA vs. AMD in OCT Scans.

| Case Number | Classifier Name | Evaluation Metrics |  |  |
| --- | --- | --- | --- | --- |
|  |  | AUC-ROC | F1 Score | Accuracy |
| 1 | ResNet50 | **$0.999 \pm 0.0$** | **$0.992 \pm 0.002$** | **$0.995 \pm 0.001$** |
| | MobileNetV2 | $0.999 \pm 0.0$ | $0.986 \pm 0.0$ | $0.992 \pm 0.0$ |
| | ViT-B/16 | $0.998 \pm 0.003$ | $0.986 \pm 0.019$ | $0.982 \pm 0.011$ |
| 2 | ResNet50 | $0.999 \pm 0.0$ | $0.979 \pm 0.006$ | $0.980 \pm 0.006$ |
|  | MobileNetV2 | **$0.999 \pm 0.0$** | **$0.994 \pm 0.003$** | **$0.994 \pm 0.003$** |
| | ViT-B/16 | $0.999 \pm 0.0$ | $0.964 \pm 0.013$ | $0.964 \pm 0.013$ |

DL = Deep Learning; AUC-ROC = Area Under the Receiver Operating Characteristic curve; GA = Geographic Atrophy; AMD = Age-related Macular Degeneration.

Interestingly, when using only B-scans of interest (Case 2), MobileNetV2 demonstrated the best overall performance with an AUC-ROC of 0.999 ± 0.0, F1 score of 0.994 ± 0.003, and accuracy of 0.994 ± 0.003. This suggests that for the task of differentiating GA from other AMD forms, a lighter architecture like MobileNetV2 can achieve optimal results when focusing on specific B-scans of interest, potentially offering computational efficiency without sacrificing diagnostic accuracy.

The ROC curves in Figure 3(e-f) confirm the exceptional performance across all models for the GA vs. AMD classification task. In both Case 1 (Figure 3e) and Case 2 (Figure 3f), the curves for all three models track almost perfectly along the top-left border of the ROC space, with AUC values of 0.999 for all models except MobileNetV2 in Case 1 (AUC = 0.998). These near-perfect ROC curves indicate that the models achieve extremely high true positive rates even at very low false positive rates, suggesting that the distinguishing features between GA and other forms of AMD are consistently and reliably captured by all three architectures.

The confusion matrices in Figure 4(e-f) further illustrate this exceptional performance. In Case 1 (Figure 4e), the ViT-B/16 model correctly classified 2174 out of 2176 AMD samples (99.9% sensitivity) and 1298 out of 1305 GA samples (99.5% specificity), with only 2 AMD samples misclassified as GA and 7 GA samples misclassified as AMD. Similar excellence was observed in Case 2 (Figure 4f), with 2096 out of 2176 AMD samples correctly identified (96.3% sensitivity) and 235 out of 236 GA samples correctly classified (99.6% specificity). These results demonstrate that the discrimination between GA and other forms of AMD represents a classification task where DL approaches excel, likely due to the distinct morphological differences visible in OCT scans.

### Multi-class Classification of CGA vs. NCGA vs. NGA

The multi-class classification task of simultaneously distinguishing CGA, NCGA, and NGA presented a greater challenge for all models, as evident in Table 5. ViT-B/16 consistently outperformed the other architectures across both experimental approaches. When using all B-scans (Case 1), ViT-B/16 achieved an AUC-ROC of 0.803 ± 0.003, F1 score of 0.593 ± 0.002, and accuracy of 0.694 ± 0.004.

**Table 5:**
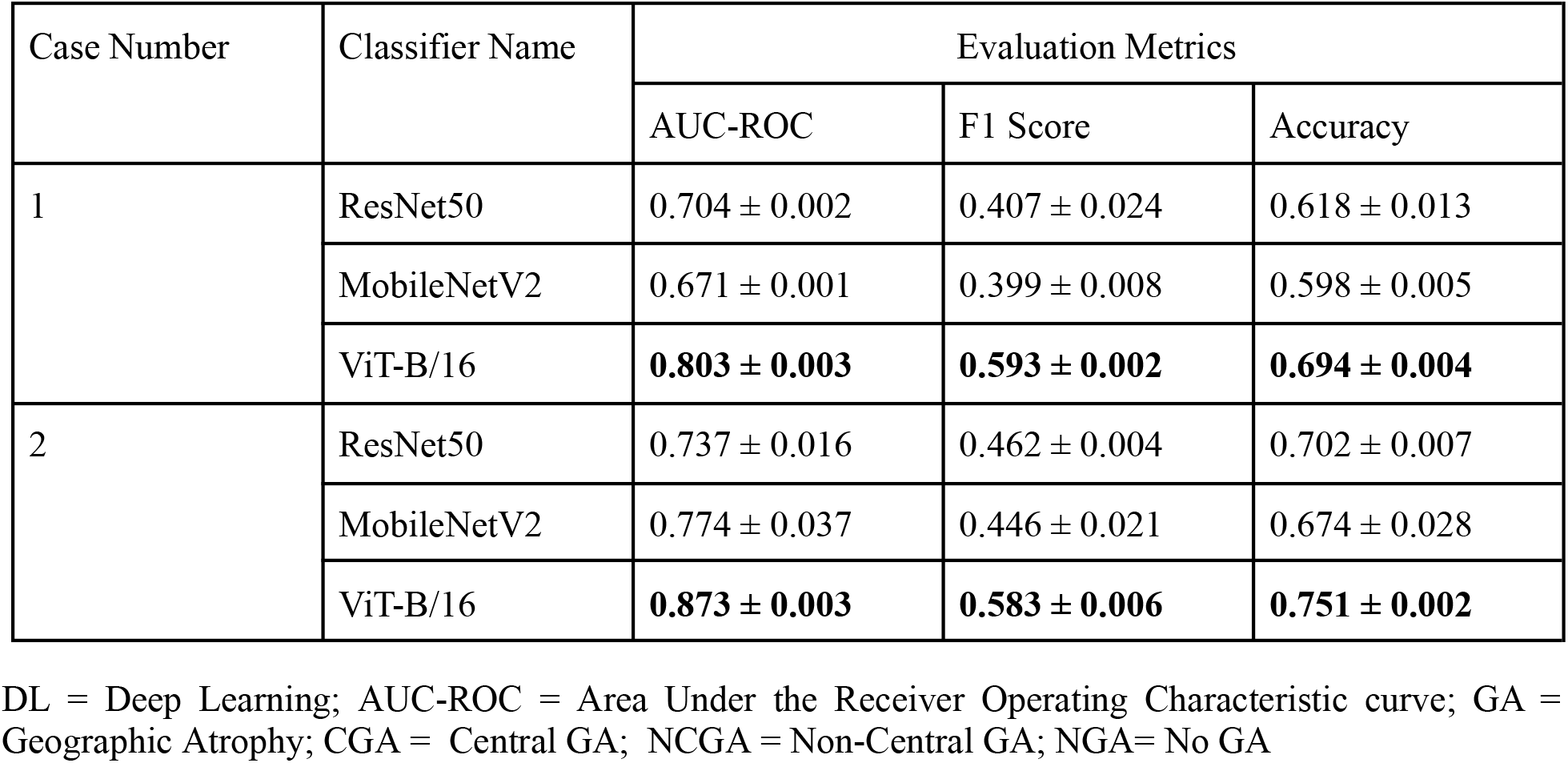
Multi-class Classification Performance of DL Models for Distinguishing CGA, NCGA, and NGA in OCT Volumes.

| Case Number | Classifier Name | Evaluation Metrics |  |  |
| --- | --- | --- | --- | --- |
|  |  | AUC-ROC | F1 Score | Accuracy |
| 1 | ResNet50 | $0.704 \pm 0.002$ | $0.407 \pm 0.024$ | $0.618 \pm 0.013$ |
| | MobileNetV2 | $0.671 \pm 0.001$ | $0.399 \pm 0.008$ | $0.598 \pm 0.005$ |
|  | ViT-B/16 | **$0.803 \pm 0.003$** | **$0.593 \pm 0.002$** | **$0.694 \pm 0.004$** |
| 2 | ResNet50 | $0.737 \pm 0.016$ | $0.462 \pm 0.004$ | $0.702 \pm 0.007$ |
| | MobileNetV2 | $0.774 \pm 0.037$ | $0.446 \pm 0.021$ | $0.674 \pm 0.028$ |
|  | ViT-B/16 | **$0.873 \pm 0.003$** | **$0.583 \pm 0.006$** | **$0.751 \pm 0.002$** |

DL = Deep Learning; AUC-ROC = Area Under the Receiver Operating Characteristic curve; GA = Geographic Atrophy; CGA = Central GA; NCGA = Non-Central GA; NGA= No GA

The performance of ViT-B/16 further improved when using only B-scans of interest (Case 2), reaching an AUC-ROC of 0.873 ± 0.003, F1 score of 0.583 ± 0.006, and accuracy of 0.751 ± 0.002. This represents an 8.7% increase in AUC-ROC and 8.2% increase in accuracy compared to using all B-scans, further supporting the efficacy of our selective B-scan approach for multi-class classification tasks.For this multi-class classification task, ViT-B/16 demonstrated statistically significant improvements over ResNet50 in both Case 1 (mean difference = 0.099, t(4) = −44.73, p < 0.001) and Case 2 (mean difference = 0.136, t(4) = −15.73, p < 0.001).

The ROC curves in Figure 3(g-h) for the multi-class classification task reflect the increased complexity of simultaneously distinguishing CGA, NCGA, and NGA. In Case 1 (Figure 3g), ViT-B/16 demonstrated the best performance with an AUC of 0.805, significantly outperforming ResNet50 (AUC = 0.713) and MobileNetV2 (AUC = 0.675). The advantage of ViT-B/16 becomes even more pronounced in Case 2 (Figure 3h), where it achieved an AUC of 0.888 compared to MobileNetV2 (AUC = 0.790) and ResNet50 (AUC = 0.755). The clear separation between the ViT-B/16 curve and those of the other models across all threshold values in both cases underscores its superior capability for this challenging multi-class task.

The confusion matrices in Figure 4(g-h) provide insight into the specific classification challenges. In Case 1 (Figure 4g), the ViT-B/16 model correctly classified 871 out of 987 CGA samples (88.2% accuracy), but struggled with NCGA, correctly identifying only 82 out of 318 samples (25.8% accuracy). NGA classification was moderate with 300 out of 487 samples correctly identified (61.6% accuracy). Notably, 138 NCGA samples were misclassified as CGA, and 98 were misclassified as NGA, highlighting the difficulty in distinguishing NCGA from both CGA and NGA. In Case 2 (Figure 4h), while overall accuracy improved, the model continued to show limitations with NCGA classification, correctly identifying only 9 out of 38 samples (23.7% accuracy), with 17 NCGA samples misclassified as CGA and 12 as NGA. However, NGA classification improved substantially to 88.7% accuracy (63 out of 71 samples), suggesting that selective B-scan analysis particularly enhances the identification of cases without GA.

Across all classification tasks, ViT-B/16 demonstrated superior overall performance, particularly in the more challenging tasks of CGA vs. NCGA differentiation and multi-class classification. Furthermore, our results consistently show that focusing on B-scans of interest that include foveal regions and areas with distinctive GA characteristics improved model performance across most metrics, while simultaneously reducing computational requirements.

## Discussion

Our study represents the first comprehensive investigation of DL-based classification of GA subtypes using OCT imaging. Safai et al. explored 12 different DL algorithms for GA segmentation using FAF images.^10^ Cluceru et al. utilized FAF topographic imaging features to predict GA growth rate using DL-based algorithms.^18^ In another recent study, Ursula et al. used DL-based analysis of OCT to quantify morphological changes of the PRs and RPE layers under pegcetacoplan therapy in GA.^19^ While most of the studies have focused primarily on GA segmentation and progression^20–23^, we have addressed the critical need for automated classification of GA phenotypes, demonstrating that accurate distinction between CGA, NCGA, and NGA is achievable through appropriate model selection and optimization.

The distinction between CGA and NCGA has significant clinical implications for visual prognosis, patient management, and therapeutic decision-making. This distinction has become increasingly important with the recent FDA approval of complement inhibitors for GA treatment^24^. Our results demonstrate that DL approaches, particularly Vision Transformer architectures, can capture the nuanced morphological differences between GA subtypes with promising accuracy (AUC-ROC of 0.728± 0.083 for ViT-B/16 in CGA vs. NCGA classification). Such automated differentiation can potentially standardize GA phenotyping in clinical practice and research settings, reducing inter-observer variability and enabling more consistent patient stratification. Moreover, the exceptional performance in GA vs. AMD classification (AUC-ROC > 0.998) demonstrates the ability to distinguish between these pathologies based on their distinctive OCT signatures which includes GA characterized by choroidal hyper transmission and RPE loss^1^, vs other AMD pathologies. This differentiation capability has substantial implications for treatment selection as therapeutics become increasingly targeted toward specific AMD phenotypes.

Vision Transformer (ViT-B/16) consistently outperformed both ResNet50 and MobileNetV2 architectures across all classification tasks, particularly for the more challenging tasks of CGA vs. NCGA differentiation and multi-class classification. We initially implemented ResNet50 as our baseline model due to its established performance in medical imaging and its proven ability to extract hierarchical features through deep residual connections^25,26^. We then explored MobileNetV2 to investigate whether a more computationally efficient architecture can maintain diagnostic accuracy while reducing processing requirements which is an important consideration for potential clinical deployment. Finally, we implemented ViT-B/16 to leverage its self-attention mechanisms that capture global spatial relationships between distant image regions^27–29^. ViT’s technical superiority stems from its ability to model long-range dependencies through self-attention mechanisms, which is crucial for GA classification where the spatial relationship between atrophic regions and the foveal center determines the GA subtype. Unlike convolutional networks with fixed receptive fields, ViT’s attention mechanism simultaneously considers both tissue characteristics and their positional relationships to the fovea, effectively connecting spatially distant but clinically related features. Additionally, our multi-class classification (CGA vs. NCGA vs. NGA) achieved 75% accuracy with ViT-B/16. This performance hierarchy aligns with clinical experience, where distinguishing NCGA from early CGA can challenge even experienced clinicians. Despite its lighter architecture ^16^, MobileNetV2 performed well in several tasks when using selective B-scans. It suggests that efficient models can achieve good results when focused on the most relevant images and that computational efficiency need not compromise diagnostic performance when the input data is optimally selected.

Our comparison between using all B-scans (Case 1) vs only diagnostically relevant B-scans containing foveal regions (Case 2) yielded important insights. The selective B-scan approach consistently improved model performance across most classification tasks, particularly for ViT-B/16 in the GA vs. NGA task (AUC-ROC improved from 0.856 to 0.950, a 11.0% increase). This finding has significant implications for clinical implementation. By focusing only on clinically relevant B-scans, we substantially reduced computational requirements without sacrificing diagnostic accuracy, which is crucial for real-time clinical applications. The improved performance with selective B-scans suggests that inclusion of non-informative B-scans may introduce noise that hinders model learning. This parallels clinical practice, where ophthalmologists typically focus on specific B-scans containing foveal structures when making GA assessments. Furthermore, the selective B-scan approach mirrors how specialists evaluate OCT volumes in practice, potentially improving clinical interpretability and adoption of these automated systems.

To better understand how each model architecture interprets OCT features during classification, we employed Gradient-weighted Class Activation Mapping (Grad-CAM) visualization as shown in Figure 5^30,31^. These visualizations reveal critical differences in how the models focus on retinal structures for classification decisions. In the heat map overlays, red and yellow areas indicate regions of highest activation (most influential for the model’s decision), green areas represent moderate importance, and blue areas indicate minimal influence on classification outcomes.Our visualization analysis reveals distinct attention mechanisms across architectures. ResNet50 and MobileNetV2 demonstrate similar broad attention patterns, focusing on extensive retinal areas with diffuse activation across multiple layers. This pattern is particularly evident in their classification of CGA samples (row 1), where both models correctly identify the central atrophy but activate across wider regions than necessary. When presented with NCGA (row 2), both convolutional models misclassify it as CGA while focusing on similar structural features as in true CGA cases, indicating their inability to precisely discriminate the spatial relationship between atrophic regions and the foveal center. In contrast, ViT-B/16 exhibits more precise feature localization. For correct CGA classification (row 1), ViT focuses specifically on the RPE disruption at the foveal center. The transformer architecture’s self-attention mechanism enables it to simultaneously process global context and local details, as demonstrated by its activation pattern highlighting both the atrophic region and its spatial relationship to the foveal center. This targeted attention explains ViT’s superior performance in distinguishing CGA from NCGA. In general GA detection (row 3), all models correctly classify the sample, but with notable differences in attention patterns. The convolutional models display gradient-based activations that spread across retinal layers, while ViT shows discrete, point-specific attention at the RPE-Bruch’s membrane complex where GA pathology is most evident. The most revealing insights come from misclassification cases (rows 2 and 5). When ResNet50 and MobileNetV2 misclassify NCGA as CGA (row 2), they focus on similar regions as with true CGA cases, failing to capture the critical spatial distinction. Similarly, when misclassifying NGA as GA (row 5), both models attend to areas with normal retinal morphology but similar reflectivity patterns as atrophic regions. ViT-B/16 demonstrates fewer misclassifications overall, but when errors occur, its attention maps still provide interpretable evidence of the decision process.

**Figure 5:**
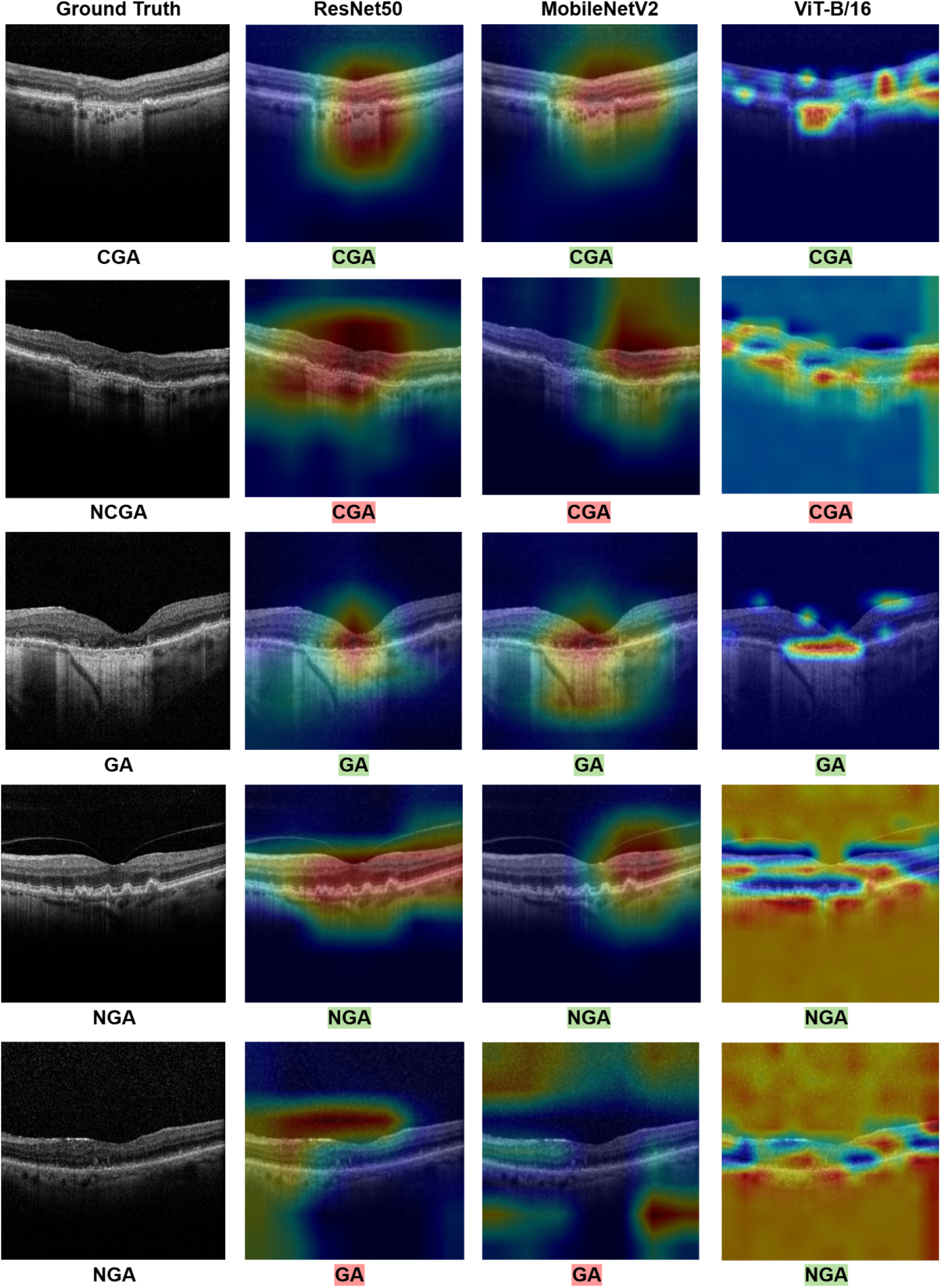
Grad-CAM visualizations revealing feature attention patterns across DL architectures for GA classification (Predictions with green highlighted text indicate correct classifications, while red text indicates misclassifications). Grad-CAM = Gradient-weighted Class Activation Mapping; DL = Deep Learning; GA = Geographic Atrophy; CGA = Central GA; NCGA = Non-Central GA; NGA = No GA; AMD = Age-related Macular Degeneration.

In conclusion, our study demonstrates that DL approaches offer promising tools for automated GA classification that can standardize phenotyping, enhance clinical trials, and serve as a clinical decision support tool. Beyond treatment selection, our automated classification system can enhance risk stratification for GA progression. Studies have shown that NCGA has variable rates of progression to CGA, with consequent impacts on visual function^6^. By accurately distinguishing these subtypes, clinicians can better predict disease trajectory, allowing for more informed patient counseling and personalized follow-up scheduling.

## Data Availability

All data produced in the present study are available upon reasonable request to the authors

## Abbreviations

AI: Artificial Intelligence
AMD: Age-related Macular Degeneration
AUC-ROC: Area Under the Receiver Operating Characteristic Curve
BCVA: Best Corrected Visual Acuity
CAM: Classification of Atrophy Meetings
GA: Geographic Atrophy
CGA: Central Geographic Atrophy
CRVO: Central Retinal Vein Occlusion
DL: Deep Learning
DME: Diabetic Macular Edema
DS1–DS4: Public OCT Datasets 1 through 4
FAF: Fundus Autofluorescence
GA: Geographic Atrophy
Grad-CAM: Gradient-weighted Class Activation Mapping
LogMAR: Logarithm of the Minimum Angle of Resolution
NCGA: Non-Central Geographic Atrophy
NGA: No Geographic Atrophy
OCT: Optical Coherence Tomography
OD: Oculus Dexter (Right Eye)
OS: Oculus Sinister (Left Eye)
PRs: Photoreceptors
RPE: Retinal Pigment Epithelium

## Declaration of generative AI in scientific writing

During the preparation of this work the authors used generative artificial intelligence tools for English language editing and grammar correction. After using this tool, the authors reviewed and edited the content as needed and took full responsibility for the content of the publication.

## Author Contributions

Sadia Siraz: Methodology, data analysis, visualization, writing - original draft.

Hindolo Kamanda: Data collection, resources.

Sina Gholami, Ahammed Sakir Nabil: Validation, writing - review & editing.

Sally Shin Yee Ong, M.D.: Supervision, validation, writing - review & editing

Minhaj Nur Alam, Ph.D.: Conceptualization, supervision, funding acquisition, writing - review & editing.

All authors read and approved the final manuscript.

## Code Availability

Codes are available at https://github.com/psydia/GA_Classification.git. The original contributions presented in the study are included in the article, further inquiries can be directed to the corresponding author.

## Data Sharing Statement

The data that support the findings of this study were obtained from Atrium Health Wake Forest Baptist and contain protected health information. The data are not publicly available due to privacy restrictions and HIPAA regulations. Requests for a de-identified, limited dataset may be considered and should be directed to the corresponding author. Any data sharing will require appropriate data use agreements and institutional approvals.

